## Supplementary information for "Evaluation of isotype specific salivary antibody assays for detecting previous SARS-CoV-2 infection in children and adults"

[1]School of Population Health Sciences, Bristol Medical School, University of Bristol, UK. [2]Bristol Veterinary School, University of Bristol, UK. [3]Bristol Vaccine Centre, School of Cellular and Molecular Medicine, University of Bristol, UK. [4]School of Biochemistry, University of Bristol, Bristol, UK. [5]BrisSynBio, University of Bristol, Bristol, UK. [6]Imophoron Ltd, Science Creates, Old Market, Midland Road, Bristol, UK.[7]School of Chemistry, University of Bristol, Bristol, UK. [8]Translational Health Sciences, Bristol Medical School, University of Bristol, UK.[9]NIHR Blood and Transplant Research Unit in Red Cell Products, University of Bristol, UK. [10]Bristol Vaccine Centre, School of Population Health Sciences, University of Bristol, UK. [11]School of Cellular and Molecular Medicine, University of Bristol, UK. [12]Bristol Bioresource Laboratories, Population Health Sciences, Bristol Medical School, University of Bristol, UK. [13]Bristol Royal Hospital for Children, University Hospitals Bristol and Weston NHS Foundation Trust, Upper Maudlin Street, Bristol, BS2 8BJ. [14]NIHR Bristol Biomedical Research Centre, University of Bristol, UK. [15]Hospital Pediátrico, Centro Hospitalar e Universitário de Coimbra, Portugal. Faculdade de Medicina, Universidade de Coimbra. [16]MRC Integrative Epidemiology Unit, Population Health Sciences, Bristol Medical School, Bristol, UK. [17]Max Planck Bristol Centre for Minimal Biology, University of Bristol, Bristol, UK. [18]Paediatric Immunology & Infectious Diseases, Bristol Royal Hospital for Children, Bristol, UK.

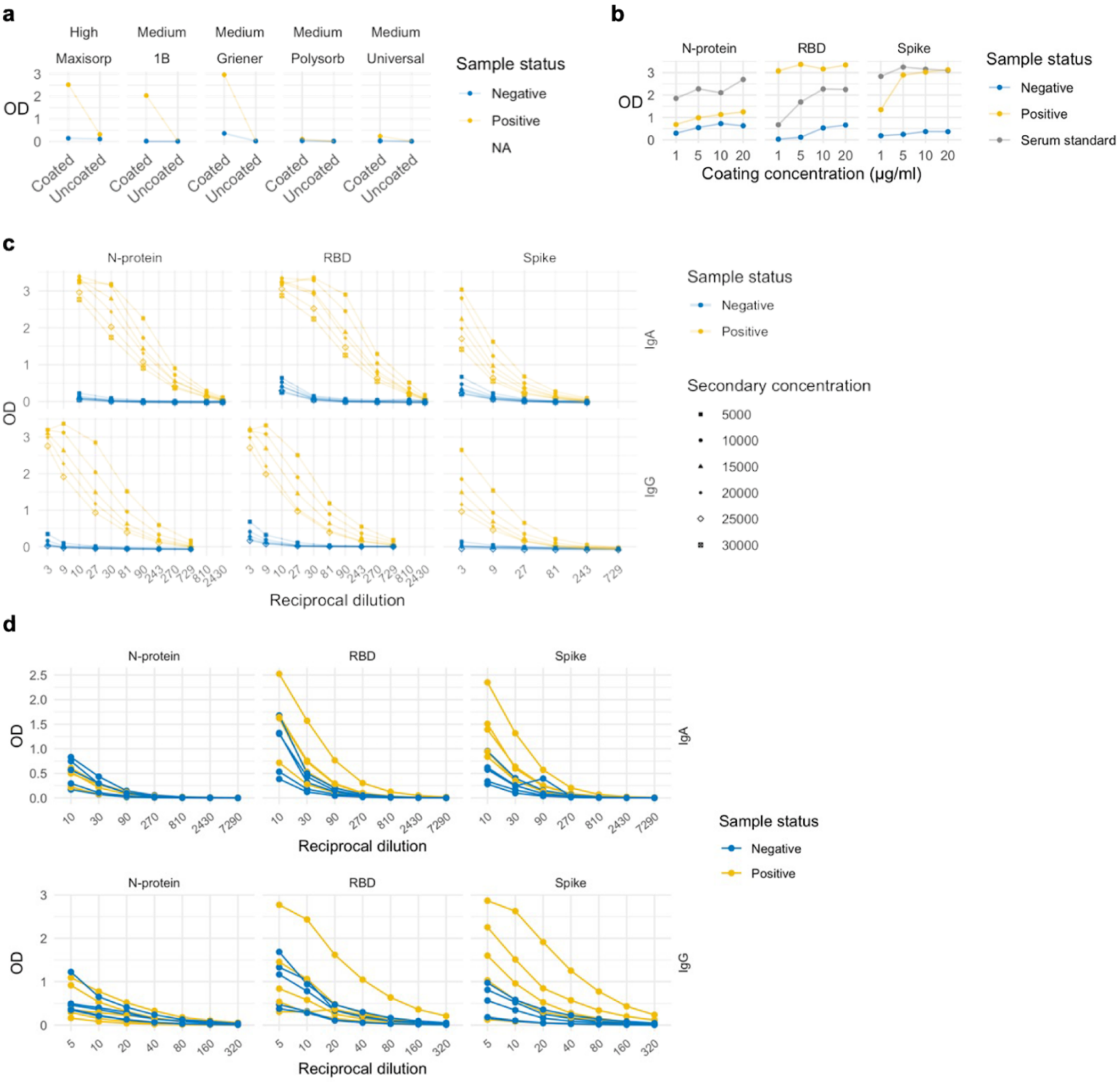

**Figure S1: Optimisation of SARS-CoV-2 salivary ELISA conditions. a,** Comparison of binding to high and medium binding plates based on reactivity to spike IgA. Negative sample: no previous SARS-CoV-2 exposure history or symptoms; positive sample from a clinically suspected donor (laboratory unconfirmed infection) **b,** Optimisation of antigen coating concentration based on responses to N-protein, RBD and Spike IgA. Blue data points show data from negative samples (healthy donor with no previous SARS-CoV-2 exposure history or symptoms); yellow data points show data from positive samples (N-protein: convalescent PCR-confirmed COVID-19 pool from 3 donors; RBD and spike: one clinically suspected COVID-19 donor); grey data points show data from serum standard (convalescent PCR-confirmed COVID-19 pool from 3 donors). **c,** Optimisation of secondary antibody (IgA and IgG) concentration and sample dilution using checkerboard titrations. Blue data points show data from negative samples (healthy donor with no previous SARS-CoV-2 exposure history or symptoms), yellow data points show data from positive samples (N-protein: convalescent PCR confirmed COVID-19 pool from 3 donors; RBD and spike IgA: one clinically suspected COVID-19 sample; RBD and spike IgG: convalescent PCR confirmed COVID-19 donor). Different shapes show data from each of the secondary antibody concentrations tested, ranging from 1 in 5,000 to 1 in 30,000. **d,** Representative serial dilution curves showing isotype specific (IgA and IgG) salivary responses to spike, RBD and N-protein. Blue data points show data from negative (pre-pandemic) samples, yellow data points show data from positive (convalescent PCR confirmed) samples.

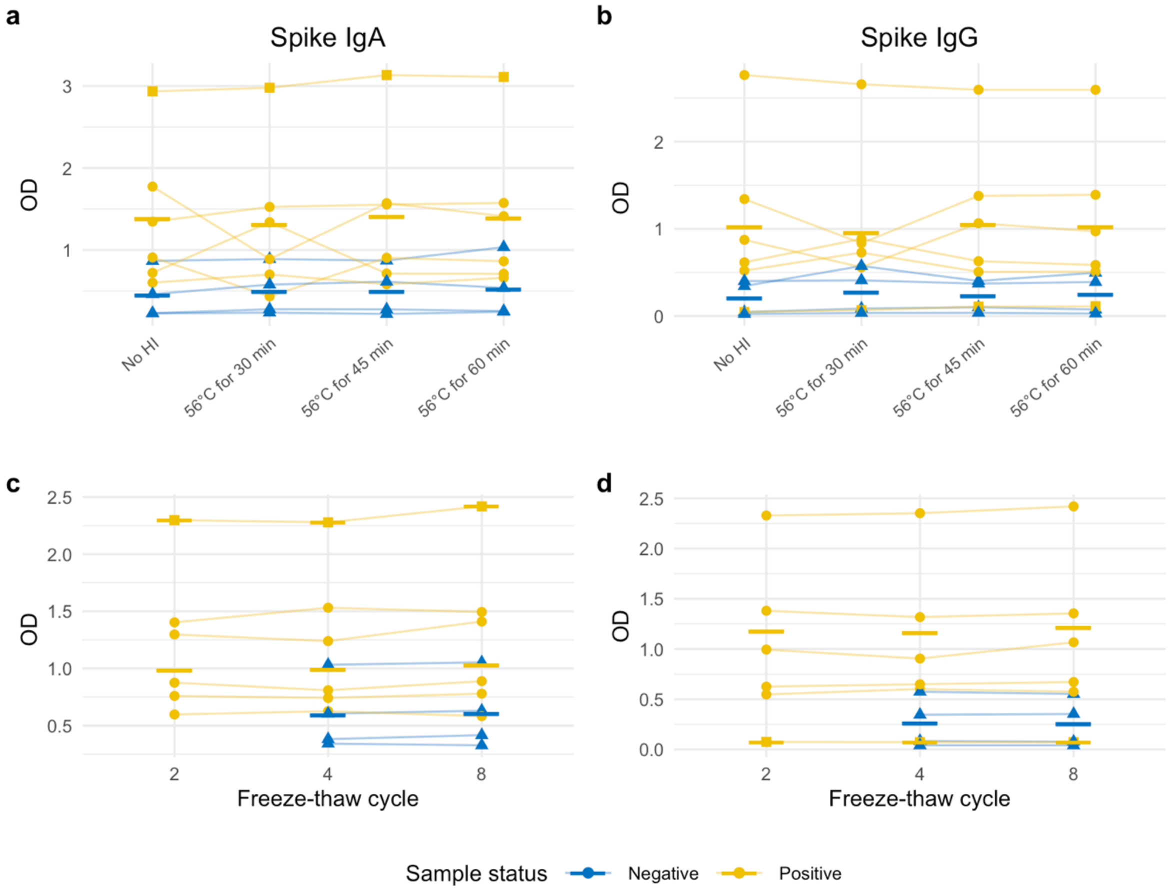

**Figure S2: Impact of heat inactivation and freeze-thawing on isotype specific reactivity to spike protein. a, c,** Reactivity to spike IgA. **b, d,** Reactivity to spike IgG. Blue triangle data points show data from negative samples (pre-pandemic), yellow circle and square data points show data from positive samples (convalescent PCR confirmed COVID-19 = circle; one clinically suspected COVID-19 sample = square. Solid bar is the mean of each negative and positive group at each condition. OD = optical density (absorbance at 450 nm and background at 570 nm). Experiments were performed once.

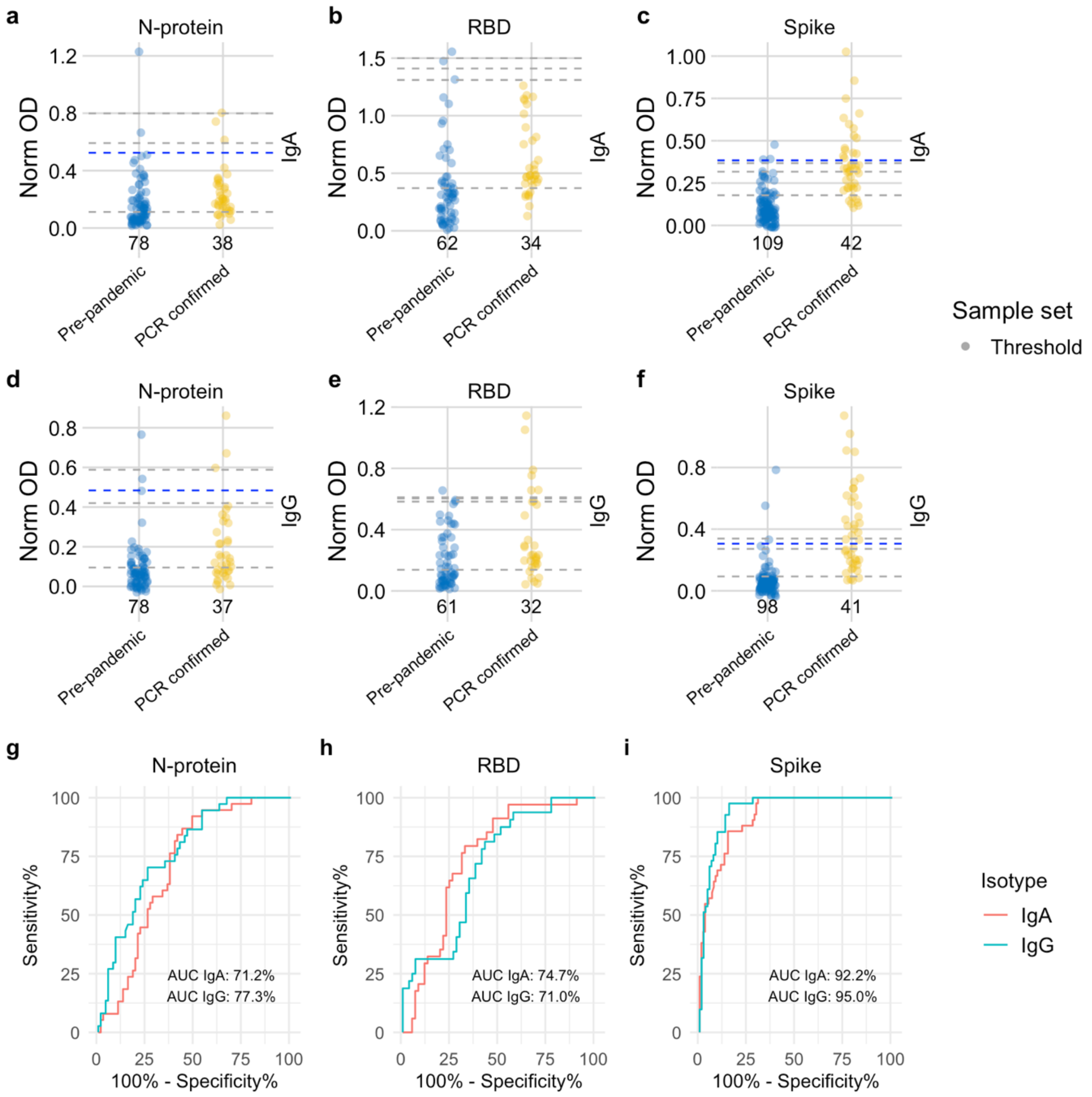

**Figure S3: Dotplots and ROC curves for threshold set samples. a-f,** Dotplots show the scatter of values. Data are presented for known negative and known positive samples tested in the threshold set, the number of samples tested in each group is shown under the corresponding data points. Proposed thresholds ($97^{th}$ – $99^{th}$ percentile and Youden's index) are shown as dashed lines. For N-protein and Spike, final thresholds are shown as dashed blue lines. **g-i**, ROC curves represent assay performance using the samples true known negative and positive status. ROC = receiver operating characteristic. AUC = area under the ROC curve. Norm OD = Normalised OD (a relative ratio to the serum standard). N-protein = Nucleocapsid protein. RBD = Receptor binding domain.

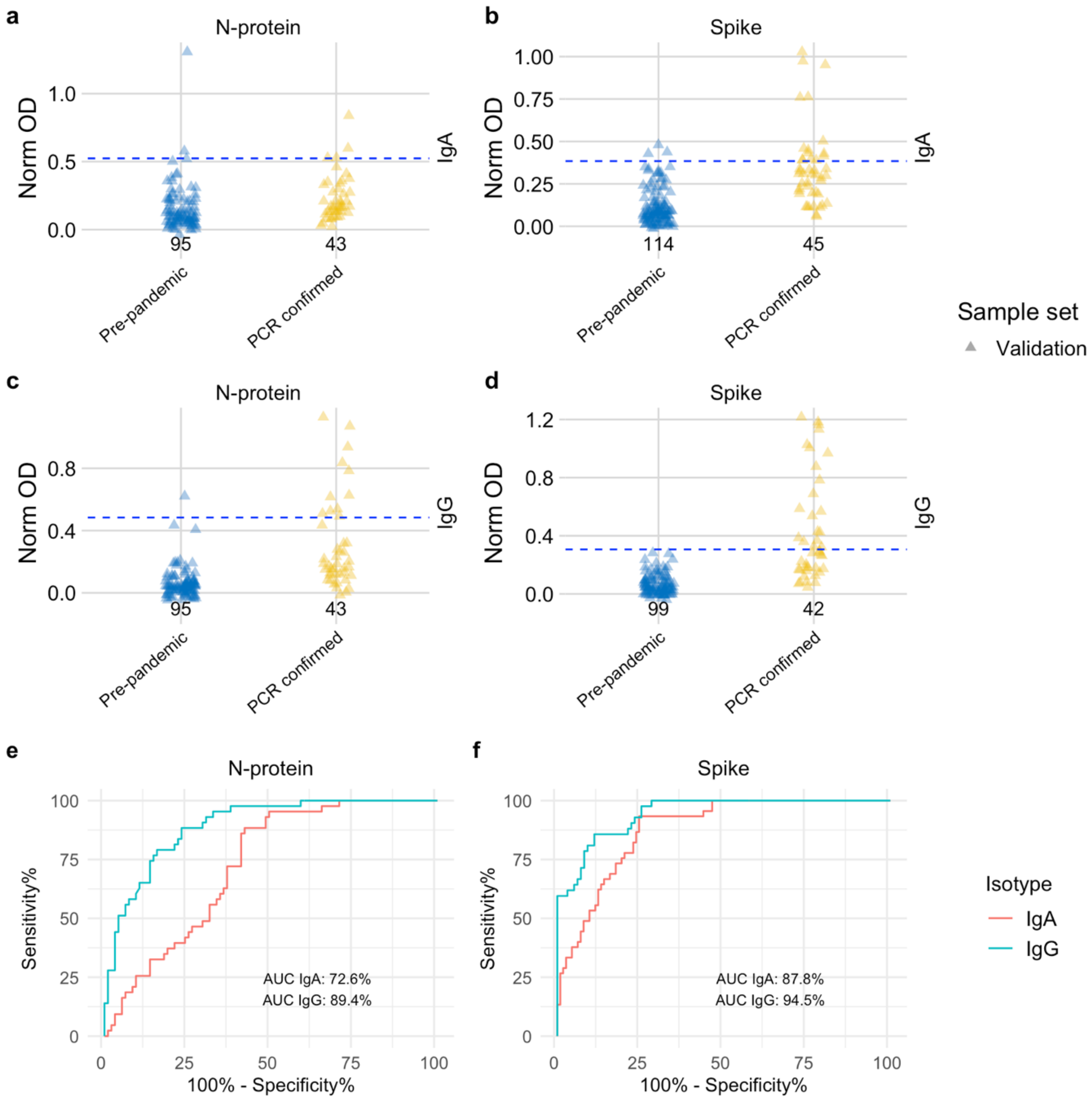

**Figure S4: Dot plots and ROC curves for validation set samples. a-d,** Dotplots show the scatter of values. Data are presented for known negative and known positive samples tested in the validation set, the number of samples tested in each group is shown under the corresponding data points. For N-protein and Spike, final thresholds are shown as dashed blue lines; **e, f**, ROC curves represent assay performance. ROC = receiver operating characteristic. AUC = area under the ROC curve. Norm OD = Normalised OD (a relative ratio to the serum standard). N-protein = Nucleocapsid protein. RBD = Receptor binding domain.

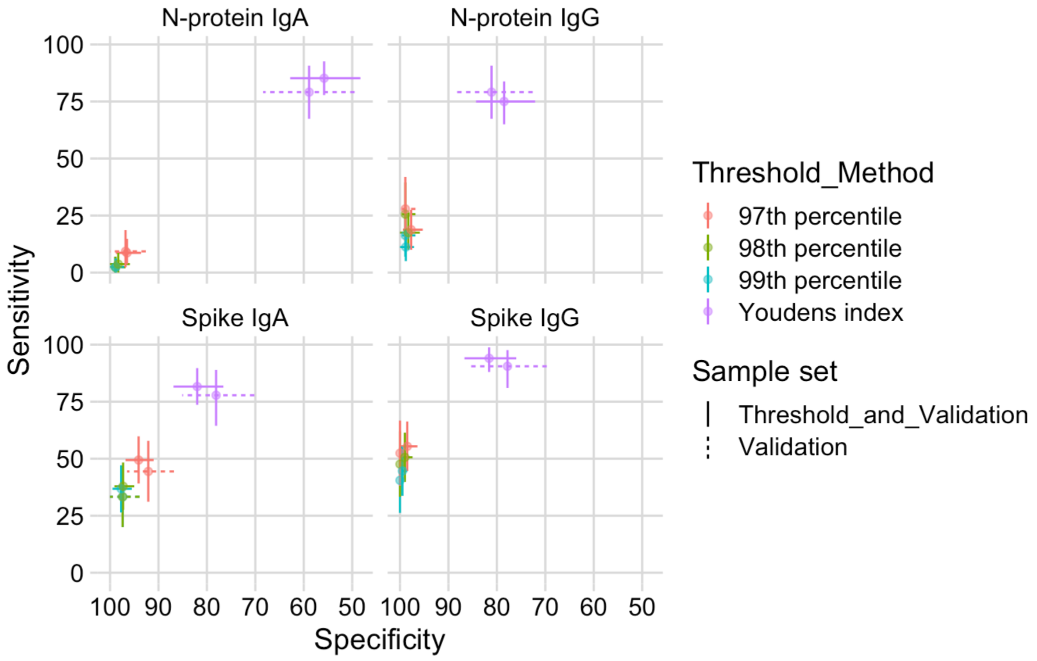

**Figure S5: Specificity and sensitivity comparison using different threshold methods.** Thresholds were applied to validation sample set only and threshold and validation sets combined. The threshold method is represented by colour and sample set by line type.

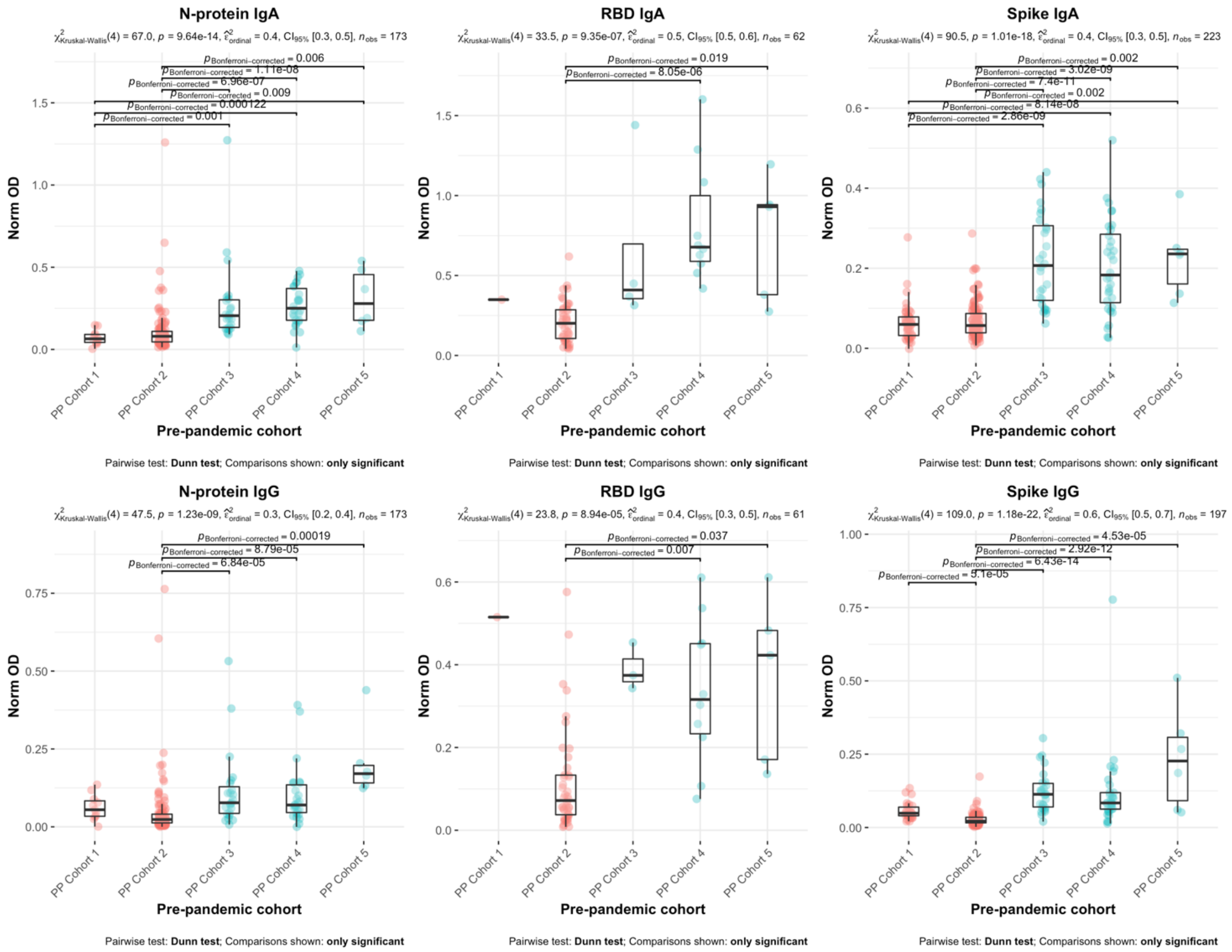

**Figure S6: SARS-CoV-2 specific IgA and IgG responses among samples collected from different pre-pandemic cohorts.** Responses stratified by age (adult vs child), represented by colour: pink = children and blue = adults. Results expressed as normalised optical density (OD); a relative ratio to the serum standard. N-protein = nucleocapsid protein; RBD = receptor binding domain. Antibody responses were compared across multiple groups using the Kruskal-Wallis test with post-hoc testing using Dunn's test. Significant Bonferroni corrected pair-wise comparisons are shown (p≤0.05).

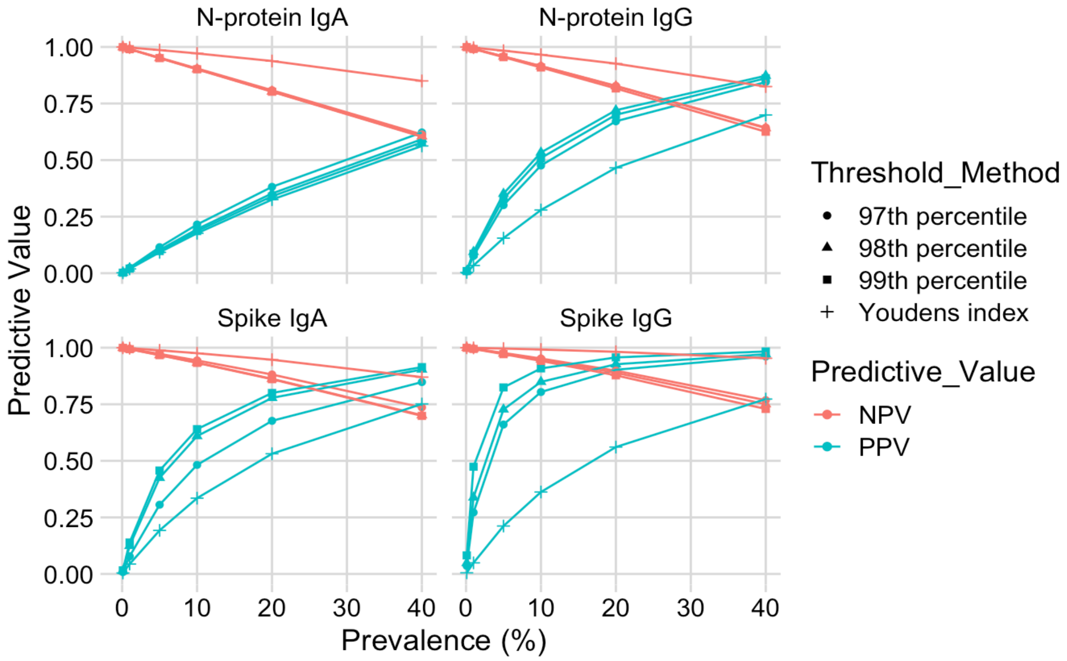

**Figure S7: Positive and negative predictive values for N-protein IgA and IgG and Spike IgA and IgG assays over 4 different thresholds.** These are implied values of PPV (positive predictive value) and NPV (negative predictive value) based on Table 2 sensitivity and specificity estimates. NPVs are shown in pink and PPVs are shown in blue. Each of the different threshold methods are shown with a different shape: 97$^{th}$ percentile = circle; 98$^{th}$ percentile = triangle; 99$^{th}$ percentile = square; Youden's index = cross.

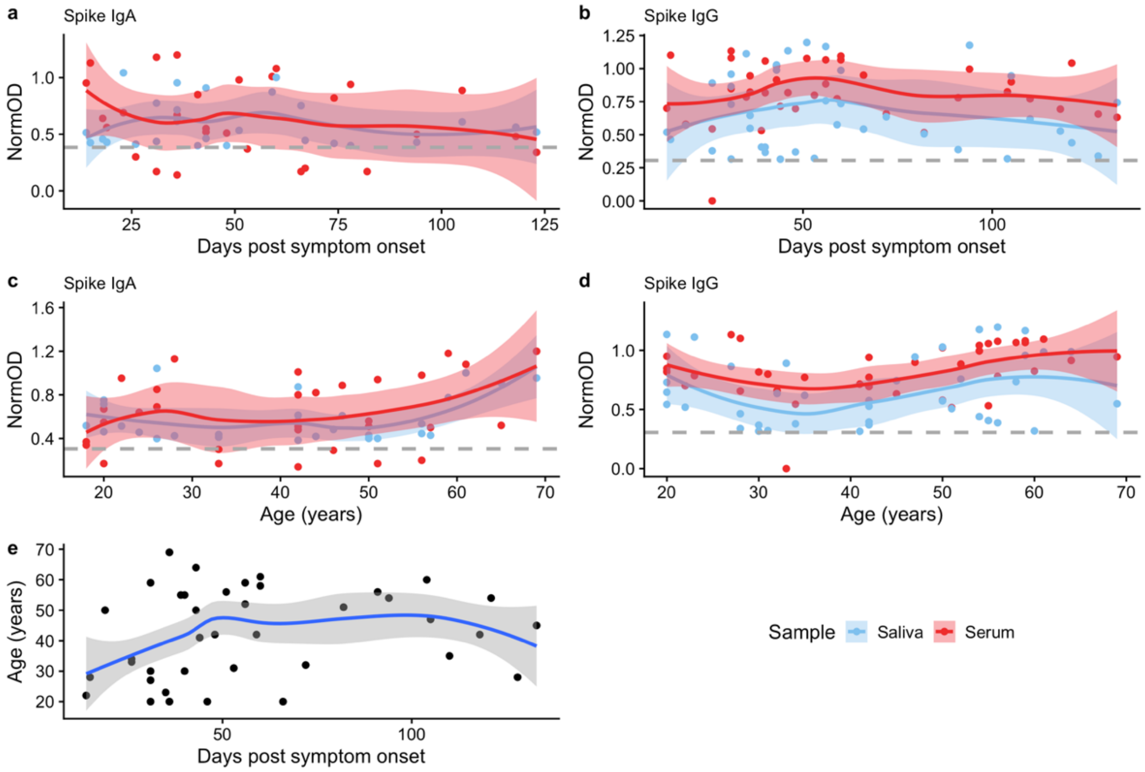

**Figure S8: Dynamics of salivary and serum spike antibody responses. a-d,** Distribution of salivary and serum spike IgA and IgG antibody in relation to time since symptom onset and age. Salivary samples positive for either spike IgA (n=28) or spike IgG (n=40) according to validated thresholds (shown as grey dotted lines) and corresponding paired serum samples are shown. **e,** Relationship between age of participant and days post symptom onset. Each point is an individual, lines and shaded areas are representative of the smoothed mean (loess method) and corresponding 95% confidence interval. Grey dashed horizontal line indicates the thresholds for salivary spike IgA and IgG positivity. NormOD = normalised OD (relative ratio to a pooled serum standard).

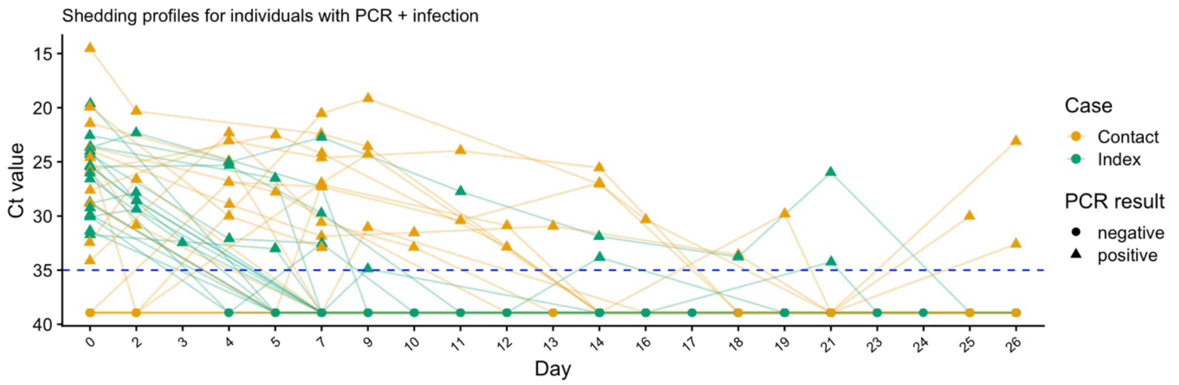

**Figure S9: Shedding profiles for PCR-confirmed index cases (n=19) and PCR-confirmed contacts (n=17) during the household study.** Saliva samples were tested by RT-qPCR for SARS-CoV-2 infection. The threshold for positivity was 35 cycles, shown as the horizontal blue dashed line. Contacts and index cases are shown in yellow and green respectively. Shape denotes PCR result: negative = circle; positive = triangle.

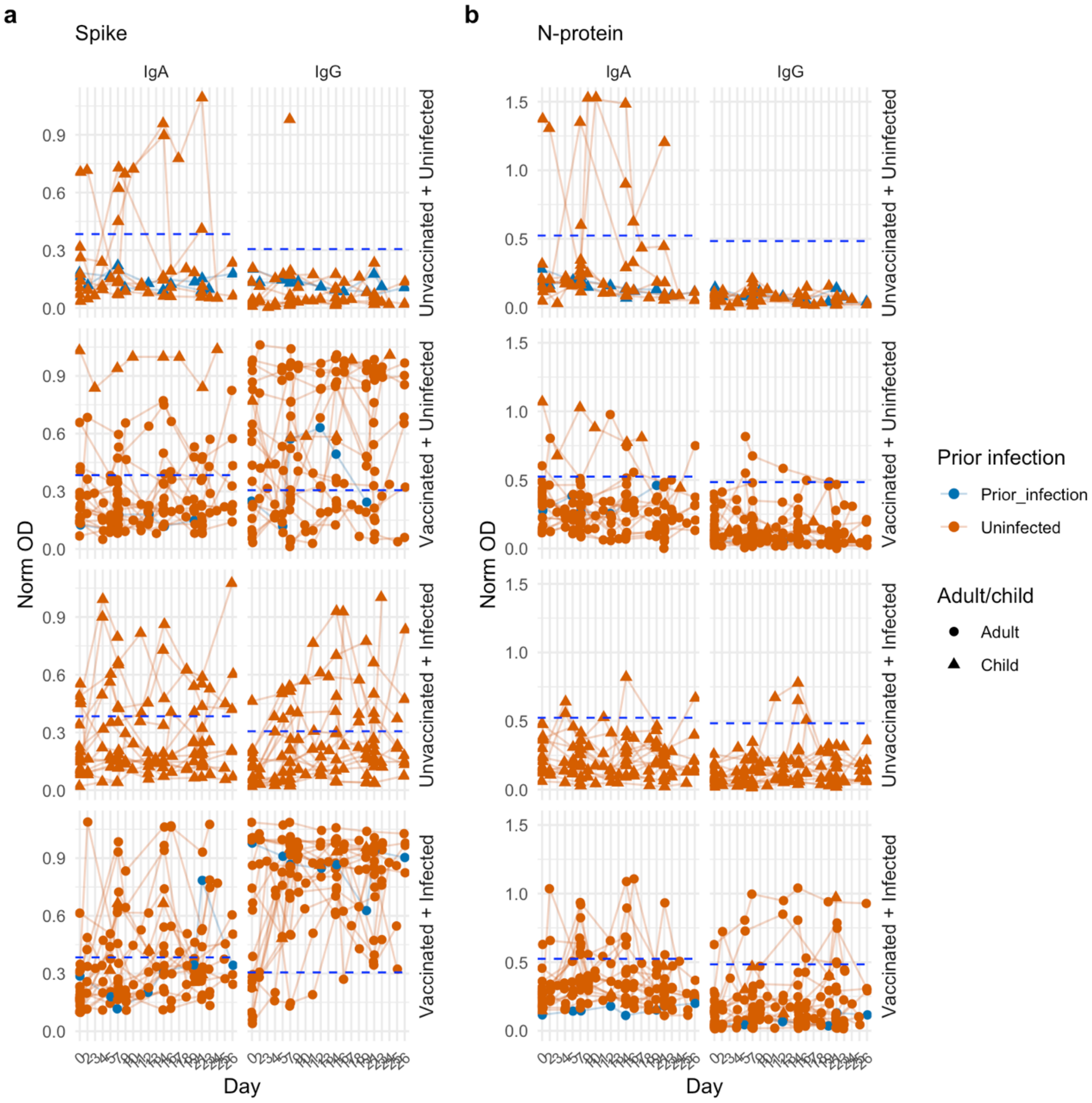

**Figure S10: Dynamics of salivary N-protein and spike IgA and IgG responses in children and adults in household outbreaks.** Responses stratified by COVID-19 vaccination, infection status based on RT-qPCR performed on the same saliva sample tested for antibodies and participant (adult/child status). Top two panels show antibody responses for uninfected individuals by vaccination status. Bottom two panels show antibody responses infected individuals by vaccination status. Samples from adults are represented with a circle, samples from children (<18 years) are represented with a triangle. Four individuals with infection ≥10 days prior to Day 0 are shown in blue. Thresholds for antibody positivity are indicated with a blue dashed horizontal line. Norm OD = Normalised OD (a relative ratio to the serum standard). N-protein = Nucleocapsid protein.

**Table S1: Evaluation of threshold methods.** Using samples in the threshold set, performance over four thresholds was evaluated. AUC = Area under the curve. 95% CI = 95% confidence interval; calculated for AUC using DeLong's method or computed with 10,000 stratified bootstrap replicates for sensitivity and specificity estimates.

| | | | Threshold sample set | | | |
|---|---|---|---|---|---|---|
| **Assay** | **n pre-pandemic** | **n PCR-confirmed** | **AUC** | | **95% CI** | |
| **Spike IgA** | 109 | 42 | 92.2 | | 88.1-96.3 | |
| **Spike IgG** | 98 | 41 | 94.9 | | 91.6-98.3 | |
| **N-protein IgA** | 78 | 38 | 71.2 | | 62.0-80.5 | |
| **N-protein IgG** | 78 | 37 | 77.3 | | 68.7-85.9 | |
| **RBD IgA** | 62 | 34 | 74.7 | | 64.8-84.6 | |
| **RBD IgG** | 61 | 32 | 70.6 | | 60.0-81.2 | |
| | **Threshold method** | **Threshold value (Normalised OD)** | **Specificity** | **95% CI** | **Sensitivity** | **95% CI** |
| **Spike IgA** | 99th percentile | 0.384 | 98.2 | 95.4-100 | 40.5 | 26.2-54.8 |
| | 98th percentile | 0.368 | 97.2 | 93.6-100 | 42.9 | 28.6-57.1 |
| | 97th percentile | 0.317 | 96.3 | 92.7-99.1 | 54.8 | 40.5-69 |
| | Youden's index | 0.178 | 85.3 | 78.9-91.7 | 85.7 | 73.8-95.2 |
| **Spike IgG** | 99th percentile | 0.339 | 98 | 94.9-100 | 48.8 | 34.1-63.4 |
| | 98th percentile | 0.306 | 96.9 | 92.9-100 | 53.7 | 39-68.3 |
| | 97th percentile | 0.272 | 95.9 | 91.8-99 | 58.5 | 43.9-73.2 |
| | Youden's index | 0.093 | 84.7 | 77.6-91.8 | 97.6 | 92.7-100 |
| **N-protein IgA** | 99th percentile | 0.799 | 98.7 | 96.2-100 | 2.63 | 0-7.89 |
| | 98th percentile | 0.592 | 97.4 | 93.6-100 | 5.26 | 0-13.2 |
| | 97th percentile | 0.524 | 96.2 | 91-100 | 7.89 | 0-18.4 |
| | Youden's index | 0.112 | 51.3 | 39.7-61.5 | 92.1 | 84.2-100 |
| **N-protein IgG** | 99th percentile | 0.588 | 98.7 | 96.2-100 | 5.41 | 0-13.5 |
| | 98th percentile | 0.484 | 97.4 | 93.6-100 | 8.11 | 0-18.9 |
| | 97th percentile | 0.420 | 96.2 | 91-100 | 8.11 | 0-18.9 |
| | Youden's index | 0.095 | 74.4 | 64.1-83.3 | 70.3 | 56.7-83.8 |
| **RBD IgA** | 99th percentile | 1.50 | 98.4 | 95.2-100 | 0 | 0-0 |
| | 98th percentile | 1.41 | 96.8 | 91.9-100 | 0 | 0-0 |
| | 97th percentile | 1.31 | 96.8 | 91.9-100 | 0 | 0-0 |
| | Youden's index | 0.371 | 67.7 | 56.5-79 | 79.4 | 64.7-91.2 |
| **RBD IgG** | 99th percentile | 0.611 | 98.4 | 95.1-100 | 18.2 | 6.1-30.3 |
| | 98th percentile | 0.604 | 96.7 | 91.8-100 | 18.2 | 6.1-30.3 |
| | 97th percentile | 0.583 | 96.7 | 91.8-100 | 18.2 | 6.1-30.3 |
| | Youden's index | 0.138 | 57.4 | 44.3 | 81.8 | 66.7-93.9 |

**Table S2: N-protein intra- and inter-assay variation. Assay variation was calculated for 10 plates assayed when testing saliva samples collected from household outbreaks.** %CV was calculated using normalised OD values for Saliva Neg† and Pos†† QCs; raw OD values were taken for Pooled + serum††† %CVs.

| Assay | Sample ID* | Mean/SD/ %CV | Plate 1 | Plate 2 | Plate 3 | Plate 4 | Plate 5 | Plate 6 | Plate 7 | Plate 8 | Plate 9 | Plate 10 | Inter-assay mean | Inter-assay SD | Inter-assay %CV |
|---|---|---|---|---|---|---|---|---|---|---|---|---|---|---|---|
| N-protein IgA | Saliva Neg QC | Mean (norm.) | 0.11 | 0.13 | 0.09 | 0.12 | 0.11 | 0.10 | 0.10 | 0.16 | 0.11 | 0.14 | 0.12 | 0.02 | 18.93 |
| | | SD | 0.02 | 0.02 | 0.01 | 0.01 | 0.02 | 0.01 | 0.02 | 0.02 | 0.01 | 0.00 | | | |
| | | %CV | 10.88 | 10.02 | 6.77 | 5.60 | 8.20 | 4.70 | 10.20 | 9.10 | 4.20 | 2.00 | | | |
| | Pooled + serum | Mean (OD) | 1.48 | 1.49 | 1.43 | 2.30 | 2.20 | 2.21 | 2.18 | 1.43 | 1.34 | 1.49 | 1.75 | 0.41 | 23.10 |
| | | SD | 0.01 | 0.00 | 0.03 | 0.05 | 0.04 | 0.02 | 0.05 | 0.03 | 0.04 | 0.01 | | | |
| | | %CV | 0.57 | 0.33 | 2.02 | 2.20 | 1.80 | 0.80 | 2.30 | 2.30 | 2.70 | 0.70 | | | |
| N-protein IgG | Saliva Neg QC | Mean (norm.) | 0.09 | 0.09 | 0.11 | 0.11 | 0.14 | 0.14 | 0.13 | 0.13 | 0.15 | 0.15 | 0.12 | 0.02 | 19.28 |
| | | SD | 0.02 | 0.01 | 0.01 | 0.00 | 0.00 | 0.02 | 0.01 | 0.01 | 0.05 | 0.01 | | | |
| | | %CV | 7.54 | 3.86 | 5.33 | 0.40 | 1.10 | 5.30 | 3.60 | 3.60 | 13.30 | 1.90 | | | |
| | Pooled + serum | Mean (OD) | 2.60 | 2.59 | 2.60 | 2.65 | 2.67 | 2.65 | 2.69 | 2.45 | 2.43 | 2.55 | 2.59 | 0.09 | 3.41 |
| | | SD | 0.02 | 0.01 | 0.01 | 0.01 | 0.03 | 0.00 | 0.00 | 0.01 | 0.01 | 0.02 | | | |
| | | %CV | 0.60 | 0.55 | 0.35 | 0.30 | 1.00 | 0.20 | 0.09 | 0.30 | 0.30 | 0.60 | | | |

* Top dilution for each standard considered: 1/10 for saliva IgA; 1/5 for saliva IgG; 1/50 for serum IgA; 1/1000 for serum IgG.

† Saliva Neg QC: used for monitoring assay performance and variation; composed of saliva from 2 individuals with no known history of COVID-19 infection.

†† Saliva Pos QC: used for monitoring assay performance and variation; composed of saliva from 3 individuals with PCR-confirmed COVID-19 infection.

†††Pooled + serum: used for standardisation of measurements across assay plates; composed of sera from 3 individuals with PCR-confirmed COVID-19 infection.

**Table S3: Spike intra- and inter-assay variation.** Assay variation was calculated for 10 plates assayed when testing saliva samples collected from household outbreaks. %CV was calculated using normalised OD values for Saliva Neg[†] and Pos[††] QCs; raw OD values were taken for Pooled + serum[†††] %CVs.

| Assay | Sample ID* | Mean/SD/ %CV | Plate 1 | Plate 2 | Plate 3 | Plate 4 | Plate 5 | Plate 6 | Plate 7 | Plate 8 | Plate 9 | Plate 10 | Inter-assay mean | Inter-assay SD | Inter-assay %CV |
|---|---|---|---|---|---|---|---|---|---|---|---|---|---|---|---|
| Spike IgA | Saliva Pos QC | Mean (norm.) | 0.72 | 0.70 | 0.80 | 0.85 | 0.84 | 0.79 | 0.80 | 0.79 | 0.81 | 0.81 | 0.79 | 0.05 | 5.95 |
| | | SD | 0.04 | 0.01 | 0.00 | 0.04 | 0.07 | 0.02 | 0.01 | 0.00 | 0.02 | 0.02 | | | |
| | | %CV | 2.37 | 0.31 | 0.00 | 1.80 | 3.80 | 0.80 | 0.50 | 0.01 | 1.10 | 0.80 | | | |
| | Saliva Neg QC | Mean (norm.) | 0.04 | 0.03 | 0.04 | 0.05 | 0.04 | 0.05 | 0.05 | 0.05 | 0.05 | 0.05 | 0.04 | 0.01 | 16.92 |
| | | SD | 0.00 | 0.00 | 0.00 | 0.00 | 0.01 | 0.01 | 0.00 | 0.01 | 0.00 | 0.01 | | | |
| | | %CV | 4.42 | 5.24 | 3.11 | 3.30 | 11.10 | 4.10 | 0.60 | 5.50 | 0.90 | 5.00 | | | |
| | Pooled + serum | Mean (OD) | 2.60 | 2.59 | 2.59 | 2.44 | 2.47 | 2.53 | 2.50 | 2.48 | 2.43 | 2.50 | 2.51 | 0.06 | 2.47 |
| | | SD | 0.02 | 0.01 | 0.00 | 0.01 | 0.19 | 0.02 | 0.02 | 0.05 | 0.00 | 0.00 | | | |
| | | %CV | 0.82 | 0.46 | 0.11 | 0.50 | 0.82 | 0.90 | 0.90 | 1.90 | 0.06 | 0.10 | | | |
| Spike IgG | Saliva Pos QC | Mean (norm.) | 0.69 | 0.58 | 0.74 | 0.77 | / | 0.77 | 0.75 | 0.76 | 0.72 | 0.76 | 0.73 | 0.06 | 8.62 |
| | | SD | 0.02 | 0.02 | 0.01 | 0.01 | / | 0.01 | 0.07 | 0.10 | 0.03 | 0.00 | | | |
| | | %CV | 1.27 | 1.29 | 0.77 | 0.60 | / | 0.30 | 3.50 | 5.00 | 1.40 | 0.20 | | | |
| | Saliva Neg QC | Mean (norm.) | 0.05 | 0.06 | 0.06 | 0.13 | / | 0.11 | 0.10 | 0.12 | 0.10 | 0.11 | 0.09 | 0.03 | 30.64 |
| | | SD | 0.00 | 0.04 | 0.00 | 0.00 | / | 0.01 | 0.01 | 0.02 | 0.01 | 0.01 | | | |
| | | %CV | 0.00 | 24.90 | 1.02 | 1.10 | / | 2.60 | 2.20 | 5.80 | 4.00 | 2.30 | | | |
| | Pooled + serum | Mean (OD) | 2.43 | 2.48 | 2.50 | 2.67 | 2.72 | 2.73 | 2.70 | 2.71 | 2.74 | 2.69 | 2.64 | 0.12 | 4.47 |
| | | SD | 0.01 | 0.04 | 0.00 | 0.02 | 0.04 | 0.01 | 0.02 | 0.03 | 0.04 | 0.02 | | | |
| | | %CV | 0.26 | 1.45 | 0.08 | 0.80 | 1.30 | 0.30 | 0.80 | 1.10 | 1.50 | 0.60 | | | |

[*] Top dilution for each standard considered: 1/10 for saliva IgA; 1/5 for saliva IgG; 1/50 for serum IgA; 1/1000 for serum IgG.

[†] Saliva Neg QC: used for monitoring assay performance and variation; composed of saliva from 2 individuals with no known history of COVID-19 infection.

[††] Saliva Pos QC: used for monitoring assay performance and variation; composed of saliva from 3 individuals with PCR-confirmed COVID-19 infections.

[†††] Pooled + serum: used for standardisation of measurements across assay plates; composed of sera from 3 individuals with PCR-confirmed COVID-19 infections.

**Table S4: Evaluation of assay performance in validation or combined sample sets.**
Performance of Spike and N-protein IgA and IgG assays was evaluated on either the validation samples only or the threshold and validation set samples combined, using normalised OD as input values. AUC = Area under the curve. 95% CI = 95% confidence interval calculated for AUC using DeLong's method or computed with 10,000 stratified bootstrap replicates for sensitivity and specificity estimates. Sp. = specificity, Sens. = sensitivity.

| | **Validation sample set** | | **Combined sample set** | |
|---|---|---|---|---|
| **Number of samples tested** | **N pre-pandemic** | **N PCR confirmed** | **N pre-pandemic** | **N PCR confirmed** |
| **Spike IgA** | 114 | 45 | 222 | 87 |
| **Spike IgG** | 99 | 42 | 196 | 83 |
| **N-protein IgA** | 95 | 43 | 172 | 81 |
| **N-protein IgG** | 95 | 43 | 172 | 80 |
| **Performance** | **AUC** | **95% CI** | **AUC** | **95% CI** |
| **Spike IgA** | 87.9 | 82.7-93.1 | 89.9 | 86.5-93.2 |
| **Spike IgG** | 94.5 | 91.2-97.9 | 95.0 | 92.8-97.3 |
| **N-protein IgA** | 72.6 | 64.3-80.9 | 71.9 | 65.7-78.1 |
| **N-protein IgG** | 89.4 | 84.2-94.7 | 84.6 | 79.9-89.4 |

| | **Threshold method** | **Sp.** | **95% CI** | **Sens.** | **95% CI** | **Sp.** | **95% CI** | **Sens.** | **95% CI** |
|---|---|---|---|---|---|---|---|---|---|
| **Spike IgA** | 99th percentile | 97.4 | 93.9-100 | 33.3 | 20-48.9 | 97.7 | 95.5-99.5 | 36.8 | 26.4-47.1 |
| | 98th percentile | 97.4 | 93.9-100 | 33.3 | 20-48.9 | 97.3 | 95-99.1 | 37.9 | 27.6-48.3 |
| | 97th percentile | 92.1 | 86.8-96.5 | 44.4 | 31.1-60 | 94.1 | 91-96.8 | 49.4 | 39.1-59.8 |
| | Youden's index | 78.1 | 70.2-85.1 | 77.8 | 66.6-88.9 | 82 | 76.6-86.9 | 81.6 | 73.6-89.7 |
| **Spike IgG** | 99th percentile | 100 | 100-100 | 40.5 | 26.2-54.8 | 99.5 | 98.5-100 | 44.6 | 33.7-55.4 |
| | 98th percentile | 100 | 100-100 | 47.6 | 33.3-61.9 | 99 | 97.4-100 | 50.6 | 39.8-61.4 |
| | 97th percentile | 100 | 100-100 | 52.4 | 38.1-66.7 | 98.5 | 96.4-100 | 55.4 | 44.6-66.3 |
| | Youden's index | 77.8 | 69.7-85.9 | 90.5 | 81-97.6 | 81.6 | 76-86.7 | 94 | 88-98.8 |
| **N-protein IgA** | 99th percentile | 98.9 | 96.8-100 | 2.3 | 0-7.0 | 98.8 | 97.1-100 | 2.47 | 0-6.17 |
| | 98th percentile | 98.9 | 96.8-100 | 2.3 | 0-7.0 | 98.3 | 95.9-100 | 3.7 | 0-8.64 |
| | 97th percentile | 98.9 | 96.8-100 | 9.3 | 2.3-18.6 | 96.5 | 93.6-98.8 | 8.64 | 3.7-14.8 |
| | Youden's index | 58.9 | 49.5-68.4 | 79.1 | 65.1-90.7 | 55.8 | 48.3-62.8 | 85.2 | 77.8-92.6 |
| **N-protein IgG** | 99th percentile | 98.9 | 96.8-100 | 16.3 | 6.98-27.9 | 98.8 | 97.1-100 | 11.2 | 5-18.8 |
| | 98th percentile | 98.9 | 96.8-100 | 25.6 | 14.0-39.5 | 98.3 | 95.9-100 | 17.5 | 10-26.2 |
| | 97th percentile | 98.9 | 96.8-100 | 27.9 | 16.3-41.9 | 97.7 | 95.3-99.4 | 18.8 | 10-27.5 |
| | Youden's index | 81.1 | 72.6-88.4 | 79.1 | 67.4-90.7 | 78.5 | 72.1-84.3 | 75 | 65-83.8 |

**Table S5: Sensitivity analysis investigating the impact of including repeated measures from the same individual on assay performance (AUC).** At each threshold (99th – 97th percentile and Youden's index) assay performance was evaluated for the full dataset (threshold and validation sample set combined with repeated measures included) and compared to the same data set with one observation per donor only using ROC curve analysis. AUC = Area under ROC curve

| | **Performance on full Dataset** | | **Performance on dataset with one observation per donor** | | **Number of samples tested in full dataset** | | **Number of samples tested in dataset with one observation per donor** | |
|---|---|---|---|---|---|---|---|---|
| **Assay** | AUC | 95% CI | AUC | 95% CI | Pre-pandemic | PCR confirmed | Pre-pandemic | PCR confirmed |
| **Spike IgA** | 89.9 | 86.5-93.2 | 91.3 | 87.6-95.3 | 222 | 87 | 108 | 79 |
| **Spike IgG** | 95.0 | 92.8-97.3 | 94.4 | 91.0-97.7 | 196 | 83 | 87 | 75 |
| **N-protein IgA** | 71.9 | 65.7-78.1 | 73.7 | 65-82.3 | 172 | 81 | 66 | 73 |
| **N-protein IgG** | 84.6 | 79.9-89.4 | 84.4 | 78.1-90.7 | 172 | 80 | 66 | 72 |

**Table S6: Sensitivity analysis investigating the impact of including repeated measures from the same individual on assay performance (specificity and sensitivity).** At each threshold (99th – 97th percentile and Youden's index) assay performance was evaluated for the full dataset (threshold and validation sample set combined with repeated measures included) and compared to the same data set with one observation per donor only. AUC = Area under curve.

| | | **Specificity and sensitivity estimates determined on full dataset** | | | | **Specificity and sensitivity estimates determined on dataset with one observation per donor** | | | |
|---|---|---|---|---|---|---|---|---|---|
| **Assay***[*] | Threshold method | Specificity | 95% CI | Sensitivity | 95% CI | Specificity | 95% CI | Sensitivity | 95% CI |
| **Spike IgA** | 99th percentile | 97.7 | 95.5-99.5 | 36.8 | 26.4-47.1 | 97.2 | 93.5-100 | 36.7 | 26.6-48.1 |
| | 98th percentile | 97.3 | 95-99.1 | 37.9 | 27.6-48.3 | 97.2 | 93.5-100 | 38 | 27.8-49.4 |
| | 97th percentile | 94.1 | 91-96.8 | 49.4 | 39.1-59.8 | 94.4 | 89.8-98.1 | 49.4 | 38-60.8 |
| | Youden's index | 82 | 76.6-86.9 | 81.6 | 73.6-89.7 | 86.1 | 79.6-92.6 | 82.3 | 73.4-89.9 |
| **Spike IgG** | 99th percentile | 99.5 | 98.5-100 | 44.6 | 33.7-55.4 | 98.9 | 96.6-100 | 44 | 33.3-56 |
| | 98th percentile | 99 | 97.4-100 | 50.6 | 39.8-61.4 | 97.7 | 94.3-100 | 48 | 37.3-60 |
| | 97th percentile | 98.5 | 96.4-100 | 55.4 | 44.6-66.3 | 97.7 | 94.3-100 | 52 | 41.3-64 |
| | Youden's index | 81.6 | 76-86.7 | 94 | 88-98.8 | 81.6 | 73.6-89.7 | 93.3 | 86.7-98.7 |
| **N-protein IgA** | 99th percentile | 98.8 | 97.1-100 | 2.47 | 0-6.17 | 100 | 100-100 | 2.74 | 0-6.85 |
| | 98th percentile | 98.3 | 95.9-100 | 3.7 | 0-8.64 | 98.5 | 95.5-100 | 4.11 | 0-9.59 |
| | 97th percentile | 96.5 | 93.6-98.8 | 8.64 | 3.7-14.8 | 95.5 | 89.4-100 | 8.22 | 2.74-15.1 |
| | Youden's index | 55.8 | 48.3-62.8 | 85.2 | 77.8-92.6 | 57.6 | 45.5-69.7 | 84.9 | 76.7-93.2 |
| **N-protein IgG** | 99th percentile | 98.8 | 97.1-100 | 11.2 | 5-18.8 | 100 | 100-100 | 12.5 | 5.56-20.8 |
| | 98th percentile | 98.3 | 95.9-100 | 17.5 | 10-26.2 | 100 | 100-100 | 19.4 | 11.1-29.2 |
| | 97th percentile | 97.7 | 95.3-99.4 | 18.8 | 10-27.5 | 98.5 | 95.5-100 | 20.8 | 12.5-30.6 |
| | Youden's index | 78.5 | 72.1-84.3 | 75 | 65-83.8 | 77.3 | 66.7-86.4 | 75 | 63.9-84.7 |

[*]Number of samples tested is given in Table S3

**Table S7: Specificity of assays and distribution of false positives by age on pre-pandemic samples.** FP = false positive.

| | | Age group (years) | | | | | | | | |
|---|---|---|---|---|---|---|---|---|---|---|
| | | **0-9** | **10-19** | **20-29** | **30-39** | **40-49** | **50-59** | **60-69** | **70+** | **Unk** |
| **N-protein IgA** | **Number of samples** | 112 | 9 | 41 | 7 | 0 | 0 | 0 | 0 | 4 |
| | **FPs** | 2 | 1 | 2 | 0 | 0 | 0 | 0 | 0 | 1 |
| | **Specificity** | 98.2% | 88.9% | 95.1% | 100.0% | NA | NA | NA | NA | 75.0% |
| **N-protein IgG** | **Number of samples** | 112 | 9 | 41 | 7 | 0 | 0 | 0 | 0 | 4 |
| | **FPs** | 2 | 0 | 1 | 0 | 0 | 0 | 0 | 0 | 0 |
| | **Specificity** | 98.2% | 100.0% | 97.6% | 100.0% | NA | NA | NA | NA | 100.0% |
| **Spike IgA** | **Number of samples** | 151 | 10 | 48 | 10 | 0 | 0 | 0 | 0 | 4 |
| | **FPs** | 0 | 0 | 4 | 0 | 0 | 0 | 0 | 0 | 1 |
| | **Specificity** | 100.0% | 100.0% | 91.7% | 100.0% | NA | NA | NA | NA | 75.0% |
| **Spike IgG** | **Number of samples** | 128 | 9 | 45 | 11 | 0 | 0 | 0 | 0 | 4 |
| | **FPs** | 0 | 0 | 0 | 2 | 0 | 0 | 0 | 0 | 1 |
| | **Specificity** | 100.0% | 100.0% | 100.0% | 81.8% | NA | NA | NA | NA | 75.0% |

**Table S8: Specificity of assays and distribution of false positives by sex on pre-pandemic samples.** FP = false positive.

| | | Sex | | |
|---|---|---|---|---|
| | | **Female** | **Male** | **Unknown** |
| **N-protein IgA** | **Number of samples** | 58 | 111 | 4 |
| | **FPs** | 1 | 4 | 1 |
| | **Specificity** | 98.3% | 96.4% | 75.0% |
| **N-protein IgG** | **Number of samples** | 58 | 111 | 4 |
| | **FPs** | 1 | 2 | 0 |
| | **Specificity** | 98.3% | 98.2% | 100.0% |
| **Spike IgA** | **Number of samples** | 80 | 139 | 4 |
| | **FPs** | 1 | 4 | 0 |
| | **Specificity** | 98.8% | 97.1% | 100.0% |
| **Spike IgG** | **Number of samples** | 67 | 124 | 4 |
| | **FPs** | 1 | 1 | 1 |
| | **Specificity** | 98.5% | 99.2% | 75.0% |

**Table S9: Sensitivity of assays and distribution of false negatives by time since symptom onset and saliva collection on PCR confirmed cases.** FN = false negative.

| | | Time since symptom onset and saliva sample | | | | |
|---|---|---|---|---|---|---|
| | | **Asymptomatic*** | **11-21 days** | **22-43 days** | **44-70 days** | **≥71 days** |
| **N-protein IgA** | **Number of samples** | 8 | 5 | 23 | 24 | 21 |
| | **FNs** | 8 | 4 | 20 | 22 | 20 |
| | **Sensitivity** | 0.0% | 20.0% | 13.0% | 8.3% | 4.8% |
| **N-protein IgG** | **Number of samples** | 8 | 5 | 23 | 23 | 21 |
| | **FNs** | 8 | 4 | 18 | 16 | 20 |
| | **Sensitivity** | 0.0% | 20.0% | 21.7% | 30.4% | 4.8% |
| **Spike IgA** | **Number of samples** | 9 | 5 | 24 | 26 | 23 |
| | **FNs** | 6 | 1 | 13 | 19 | 16 |
| | **Sensitivity** | 33.3% | 80.0% | 45.8% | 26.9% | 30.4% |
| **Spike IgG** | **Number of samples** | 9 | 5 | 23 | 24 | 22 |
| | **FNs** | 8 | 2 | 9 | 11 | 11 |
| | **Sensitivity** | 11.1% | 60.0% | 60.9% | 54.2% | 50.0% |

* Median [Min, Max] days post positive PCR test: 37 [15, 69] days

**Table S10: Performance of AdaBoost classifiers to predict positive and negative individuals using samples in the combined threshold and validation sample set.** Datasets used for model training and testing differed in size (92 – 150 individuals) depending upon the assay(s) included. See supplementary methods for full details on dataset construction.

| **Assay** | **Mean ROC AUC*** | **Standard deviation** |
|---|---|---|
| N-protein IgA | 0.539 | 0.105 |
| RBD IgA | 0.552 | 0.069 |
| RBD IgA, RBD IgG | 0.556 | 0.130 |
| N-protein IgA, N-protein IgG | 0.592 | 0.070 |
| N-protein IgG | 0.625 | 0.032 |
| RBD IgG | 0.650 | 0.178 |
| Spike IgA | 0.758 | 0.093 |
| N-protein IgA, RBD IgA, Spike IgA | 0.784 | 0.076 |
| N-protein IgA, Spike IgA | 0.786 | 0.066 |
| Spike IgG | 0.824 | 0.075 |
| Spike IgA, Spike IgG | 0.881 | 0.078 |
| N-protein IgG, Spike IgG | 0.886 | 0.044 |
| N-protein IgG, RBD IgG, Spike IgG | 0.942 | 0.070 |

* Receiver operator characteristic (ROC) curve area under curve (AUC) score obtained by performing 5-fold cross-validation. Assays ordered by mean ROC AUC score.

**Table S11: Detection of salivary antibody in household members given vaccination and infection status.** The number and proportion of individuals with antibody above thresholds for positivity on at least one occasion measured by N-protein and spike IgA and IgG assays*

| Assay | Unvaccinated PCR -ve (N=11) | Unvaccinated PCR +ve (N=16) | Vaccinated PCR -ve (N=20) | Vaccinated PCR +ve (N=20) |
|---|---|---|---|---|
| **N-protein IgA** | 4 (36.4%) | 3 (18.8%) | 7 (35.0%) | 11 (55.0%) |
| **N-protein IgG** | 0 (0%) | 2 (12.5%) | 2 (10.0%) | 4 (20.0%) |
| **N-protein IgA or IgG** | 4 (36.4%) | 4 (25.0%) | 9 (45.0%) | 12 (60.0%) |
| **Spike IgA** | 4 (36.4%) | 9 (56.3%) | 9 (45.0%) | 14 (70.0%) |
| **Spike IgG** | 1 (9.1%) | 11 (68.8%) | 17 (85.0%) | 20 (100%) |
| **Spike IgA or IgG** | 4 (36.4%) | 14 (87.5%) | 18 (90.0%) | 20 (100%) |

* All unvaccinated individuals were children. Only 2 children (adolescents aged between 16-18 years) received either 1 or 2 doses of vaccine, one of these vaccinated children became infected during the study.

### STARD Appendix

**Table S12: Study aims and objectives**

| Aims/Objectives | Description |
|---|---|
| Aim | Develop and evaluate salivary immunoassays suitable for large scale epidemiological and immunological SARS-CoV-2 studies and provide insights into mucosal immune responses to mild SARS-CoV-2 infection. |
| Objective 1 | Develop standardised salivary immunoassays for detection of anti-SARS-CoV-2 IgA and IgG antibodies in saliva to spike protein, it's RBD and N-protein |
| Objective 2 | Determine and evaluate salivary immunoassay diagnostic performance as screening assays for population salivo-prevalence surveys |
| Objective 3 | Investigate how observed salivo-antibody responses correlate with serum antibody overtime to better understand immunological profiles |
| Objective 4 | Field-test assays and characterise mucosal responses in households of adults and children with different levels of pre-existing immunity |

**Table S13: Test accuracy methods for index and reference tests**

| Method | Description |
|---|---|
| Index test | Salivary ELISA detecting IgA and IgG antibodies to SARS-CoV-2 spike protein, receptor binding domain, N-protein |
| Reference test | RT-PCR positive test performed on nose/throat swab. Selected as best available gold-standard for identifying current SARS-CoV-2 infection. |
| Test positivity index test cut-off | Test positivity cut-offs were established by setting thresholds on the pre-pandemic (known negative) population in the 'threshold set' to achieve ~98% specificity. See Table S1 for threshold values. |
| Test positivity reference test cut-off | RT-PCR tests were performed as part of national SARS-CoV-2 surveillance conducted by UK government laboratories. |

**Table S14: STARD Checklist**

| Section & Topic | No | Item | Reported on page # |
|---|---|---|---|
| **TITLE OR ABSTRACT** | | | |
| | **1** | Identification as a study of diagnostic accuracy using at least one measure of accuracy<br>(such as sensitivity, specificity, predictive values, or AUC) | Abstract |
| **ABSTRACT** | | | |
| | **2** | Structured summary of study design, methods, results, and conclusions<br>(for specific guidance, see STARD for Abstracts) | Abstract |
| **INTRODUCTION** | | | |
| | **3** | Scientific and clinical background, including the intended use and clinical role of the index test | Introduction |
| | **4** | Study objectives and hypotheses | Introduction, STARD Appendix Table S12 |
| **METHODS** | | | |
| *Study design* | **5** | Whether data collection was planned before the index test and reference standard<br>were performed (prospective study) or after (retrospective study) | Methods |
| *Participants* | **6** | Eligibility criteria | Methods |
| | **7** | On what basis potentially eligible participants were identified<br>(such as symptoms, results from previous tests, inclusion in registry) | Methods |
| | **8** | Where and when potentially eligible participants were identified (setting, location and dates) | Methods |
| | **9** | Whether participants formed a consecutive, random or convenience series | Methods |
| *Test methods* | **10a** | Index test, in sufficient detail to allow replication | Methods and STARD Appendix Table S13 |
| | **10b** | Reference standard, in sufficient detail to allow replication | Methods and STARD Appendix Table S13 |
| | **11** | Rationale for choosing the reference standard (if alternatives exist) | Methods and STARD Appendix Table S13 |
| | **12a** | Definition of and rationale for test positivity cut-offs or result categories<br>of the index test, distinguishing pre-specified from exploratory | Methods and STARD Appendix Table S13 |
| | **12b** | Definition of and rationale for test positivity cut-offs or result categories<br>of the reference standard, distinguishing pre-specified from exploratory | Methods and STARD Appendix Table S13 |
| | **13a** | Whether clinical information and reference standard results were available<br>to the performers/readers of the index test | Methods |
| | **13b** | Whether clinical information and index test results were available<br>to the assessors of the reference standard | Methods |
| *Analysis* | **14** | Methods for estimating or comparing measures of diagnostic accuracy | Methods |
| | **15** | How indeterminate index test or reference standard results were handled | Methods |
| | **16** | How missing data on the index test and reference standard were handled | Methods |
| | **17** | Any analyses of variability in diagnostic accuracy, distinguishing pre-specified from exploratory | Methods |
| | **18** | Intended sample size and how it was determined | Methods |
| **RESULTS** | | | |
| *Participants* | **19** | Flow of participants, using a diagram | Figure 1 |
| | **20** | Baseline demographic and clinical characteristics of participants | Table 1 |

| **Table S14 continued** | | | |
|---|---|---|---|
| **Section & Topic** | **No** | **Item** | **Reported on page #** |
| | **21a** | Distribution of severity of disease in those with the target condition | Table 1 |
| | **21b** | Distribution of alternative diagnoses in those without the target condition | NA |
| | **22** | Time interval and any clinical interventions between index test and reference standard | Table 1 |
| *Test results* | **23** | Cross tabulation of the index test results (or their distribution) by the results of the reference standard | Table 2 |
| | **24** | Estimates of diagnostic accuracy and their precision (such as 95% confidence intervals) | Table 2 |
| | **25** | Any adverse events from performing the index test or the reference standard | NA |
| **DISCUSSION** | | | |
| | **26** | Study limitations, including sources of potential bias, statistical uncertainty, and generalisability | Discussion |
| | **27** | Implications for practice, including the intended use and clinical role of the index test | Discussion |
| **OTHER INFORMATION** | | | |
| | **28** | Registration number and name of registry | NA |
| | **29** | Where the full study protocol can be accessed | NA |
| | **30** | Sources of funding and other support; role of funders | Acknowledgements |

### Supplementary methods

### Detection of SARS-CoV-2 infection by RT-qPCR on saliva

**Table S15: Full sequences and final concentrations of primers and probes used in the SARS-CoV-2 N6/E/MS2 RT-qPCR multiplex assay.** Probes were labelled with 6-Carboxyfluorescein (6-FAM) and Black Hole Quencher 1 (BHQ1) or Cyanine 5 (Cy5) and Black Hole Quencher 3 (BHQ3).

| Name | Position* | Sequence (5'-3') | Concentration used in RT-qPCR |
|---|---|---|---|
| CoV2_N6_F Forward Primer | 26269 | GATGCTGCTCTTGCTTTGCT | 200nM |
| CoV2_N6_R Reverse Primer | 26381 | CCTTGTTGTTGTTGGCCTTTAC | 200nM |
| CoV2_N6_P Probe (FAM) | 26332 | 6-FAM-CTGCTTGACAGATTGAACCAGCTTGAGAG-BHQ-1 | 100nM |
| E_Sarbeco_F1 Forward Primer | 2163 | ACAGGTACGTTAATAGTTAATAGCGT | 200nM |
| E_Sarbeco_R2 Reverse Primer | 2226 | ATATTGCAGCAGTACGCACACA | 200nM |
| E_Sarbeco_P1 Probe (FAM) | 2183 | 6-FAM- ACACTAGCCATCCTTACTGCGCTTCG-BHQ-1 | 100nM |
| MS2 2155 F Forward primer | 28919 | ATGGGGCACAAGTTGCAG | 50nM |
| MS2 2218 R Reverse primer | 29001 | GGGTAACGGTTGCTTGTTCA | 50nM |
| MS2 2175 P Probe (CY5) | 28940 | Cy5-TGCAGCGCCTTACAAGAAGTTCGC-BHQ-3 | 50nM |

* Position relative to SARS-CoV-2 isolate Wuhan-Hu-1 genome (accession MN908947.3) or MS2 phage genome (accession V00642.1).

### Machine learning analysis evaluating performance when combining assays

To construct a dataset, we combined the samples in the threshold and validation sample set, using readings obtained from the lowest dilution of each assay (1 in 5 for N-protein, RBD and Spike IgG; 1 in 10 for N-protein, RBD and Spike IgA). The lowest dilution was selected as little difference was observed in the separation of positive and negative individuals across the different sample dilutions tested. The resulting dataset contained entries for 318 individuals. However, only 32 individuals had readings measured for all 6 assays. To ensure we had enough data available for 5-fold cross-validation (see below), we consequently decided to limit the assay combinations we tested to those containing either the same antigen (i.e., N-protein IgG and IgA; RBD IgG and IgA; and Spike IgG and IgA) or the same secondary antibody (i.e., N-protein, RBD and Spike IgG; and N-protein, RBD and Spike IgA). Given that, owing to poor performance, the RBD assays were not taken forwards for threshold setting nor to the field studies, we also tested combining just the N-protein and Spike antigens (N-protein and Spike IgG; and N-protein and Spike IgA). This meant that the sizes of the datasets we used for model training and testing were, depending upon the assay(s) included, in the range of 92 – 150 individuals.

We trained AdaBoost classifiers to predict positive and negative individuals using the datasets described above (i.e., 13 datasets in total – 6 models were trained with the individual assays, and 7 models were trained with a combination of assays). Model performance was measured by calculating ROC AUC (area under the receiver operating characteristic curve) scores. Model training and testing were performed as part of a 5-fold cross-validation loop. Accordingly, we obtained 5 ROC AUC scores for each assay combination, which are represented as dots in Fig 3(b), whilst the mean of these 5 values is plotted as a bar. The above data analysis is available as a Jupyter notebook at https://github.com/Bristol-UNCOVER/Saliva_data_ML_analysis/blob/main/Saliva_dataset_analysis.ipynb.
The AdaBoost algorithm was imported into the notebook from the scikit-learn Python package.[1]

[1] Pedregosa, F. *et al. Scikit-learn: Machine Learning in Python*. *Journal of Machine Learning Research* vol. 12 http://scikit-learn.sourceforge.net. (2011).
